# COVID-19 deaths and hospitalizations averted by rapid vaccination rollout in the United States

**DOI:** 10.1101/2021.07.07.21260156

**Authors:** Seyed M. Moghadas, Pratha Sah, Meagan C. Fitzpatrick, Affan Shoukat, Abhishek Pandey, Thomas N. Vilches, Burton H. Singer, Eric C. Schneider, Alison P. Galvani

## Abstract

**Importance:** Randomized clinical trials have shown that the COVID-19 vaccines currently approved in the US are highly efficacious. However, more evidence is needed to understand the population-level impact of the US vaccination rollout in the face of the changing landscape of COVID-19 pandemic in the US, including variants with higher transmissibility and immune escape.

**Objective:** To quantify the population-level impact of the US vaccination campaign in averting cases, hospitalizations and deaths from December 12, 2020 to June 28, 2021.

**Design:** Age-stratified agent-based model which included transmission dynamics of the Alpha, Gamma and Delta variants in addition to the original Wuhan-1 variant.

**Setting:** Our model was calibrated to COVID-19 outbreak and vaccine rollout in the US. Model predictions were made at the country level.

**Participants:** Simulated age-stratified population representing US demographics.

**Main Outcomes and Measures:** Cases, hospitalizations and deaths averted by vaccination against COVID-19 in the US, compared to the counterfactuals of no vaccination and vaccination administered at half the actual pace.

**Results:** The swift vaccine rollout in the US curbed a potential resurgence of cases in April 2021, which would have been otherwise fuelled by the Alpha variant. Compared to the scenario without vaccines, we estimated that the actual vaccination program averted more than 26 million cases, 1.2 million hospitalizations and saved 279,000 lives. A vaccination campaign with half the actual rollout rate would have led to an additional 460,000 hospitalizations and 121,000 deaths.

**Conclusions and Relevance:** The COVID-19 vaccination campaign in the US has had an extraordinary impact on reducing disease burden despite the emergence of highly transmissible variants. These findings highlight that the pace of vaccination was essential for mitigating COVID-19 in the US, and underscore the urgent need to close the vaccine coverage gaps in communities across the country.

**Key Points:** *Question:* How effective was the United States (US) vaccination campaign in suppressing COVID-19 burden?

*Findings:* The vaccination campaign was highly effective in curbing the COVID-19 outbreak in the US. We estimated that the vaccine rollout saved over 275,000 lives and averted 1.2 million hospitalizations.

*Meaning:* The swift vaccine rollout in the US averted a remarkable number of cases, hospitalizations and deaths despite the emergence of highly transmissible variants.

---

The COVID-19 pandemic has unleashed devastating health and socioeconomic crises worldwide, causing more than 3.9 million deaths and 183 million reported infections globally^1^. The United States (US) alone has endured more than 600,000 deaths. The federal government has also developed, authorized, and delivered highly efficacious vaccines at an unprecedented pace. As of July 2, 2021, the US has administered more than 328 million doses of vaccine and 67% of adults have received at least one dose ^2^. The number of cases has fallen from over 300,000 per day at the apex of the pandemic in January 2021 to less than 20,000 per day in mid-June.

The precipitous decline in US cases is also notable given the emergence of more transmissible and immune-evading variants in recent months, including the B.1.1.7 (Alpha), P.1 (Gamma), and B.1.617.2 (Delta) variants. The Alpha variant, first identified in the United Kingdom, is 50% more contagious than the original SARS-CoV-2 strain^3^ and is associated with higher case fatality ^4^. The Gamma variant emerged in Brazil, and likely due to its higher transmissibility even relative to Alpha ^5^, came to dominate most pre-circulating variants in the US by mid-May ^6^. The rise of the Delta variant from the recent COVID-19 resurgence in India and other southeast Asian countries is quickly shifting the dynamics of SARS-CoV-2 in the US. Its transmissibility is the highest among all variants detected thus far ^7^. Ominously, the Delta variant is also associated with reduced neutralizing activity by antibodies in sera from convalescent or vaccinated individuals ^8^.

Understanding the impact of vaccination in suppressing COVID-19 burden is fundamental to informing strategies for improving vaccine uptake and future planning to mitigate disease outcomes. The individual-level efficacy and safety of authorized vaccines against the original viral variant are well established based on randomized controlled trials, exceeding 90% in preventing hospitalization and death ^9–11^. However, the population-level effectiveness of the vaccination campaign for the US population in the face of highly transmissible variants has not yet been evaluated.

To quantify the impact of vaccination on reducing COVID-19 burden in the US, we expanded our age-stratified agent-based model to include transmission dynamics of the Alpha, Gamma and Delta variants in addition to the original Wuhan-1 variant ^12^. The model parameters included the population demographics of the US, an empirically determined contact network accounting for pandemic mobility patterns, and age-specific risks of severe outcomes due to COVID-19. A two-dose vaccination strategy was implemented based on the number of daily vaccine doses administered for first and second doses in different age groups ^13^. Vaccine efficacies against infection, symptomatic and severe disease after each dose and for each variant were derived from published estimates (Appendix Table S3). The model was calibrated to reported incidence at the national level between October 1, 2020, and June 28, 2021.

With the calibrated model, we evaluated the impact of vaccine rollout by simulating epidemic trajectories under two counterfactual scenarios of no vaccination and a temporal vaccination rate reduced to half of the actual pace. For each scenario, cumulative infections, hospitalizations and deaths were compared to the observed pandemic trend in the US with vaccination.

We found that vaccination has markedly curbed the US pandemic burden (Figure 1). Compared to the scenario without any vaccines, we estimated that the actual vaccination program saved 279,052 (95% credible interval [CrI]: 235,326 – 324,220) lives along with averting 1,249,589 (95% CrI: 1,092,199 – 1,4094,85) hospitalizations from the start of vaccination campaign on December 12, 2020 to June 28, 2021 (Appendix Fig S1). The number of cases averted during the same period was estimated to exceed 26 million. Strikingly, vaccination prevented a substantial surge that would have occurred in April 2021, during which the US could have experienced 4,501 (95% CrI: 3,035 – 6,301) deaths at the peak (Appendix, Fig S2). This toll would have exceeded the peak during the 2020-2021 winter surge due to the higher case fatality associated with the Alpha variant relative to the original virus. Instead, due to the prioritization of elderly individuals for vaccination, overall case fatality dropped despite the dominance of Alpha.

**Figure 1.**
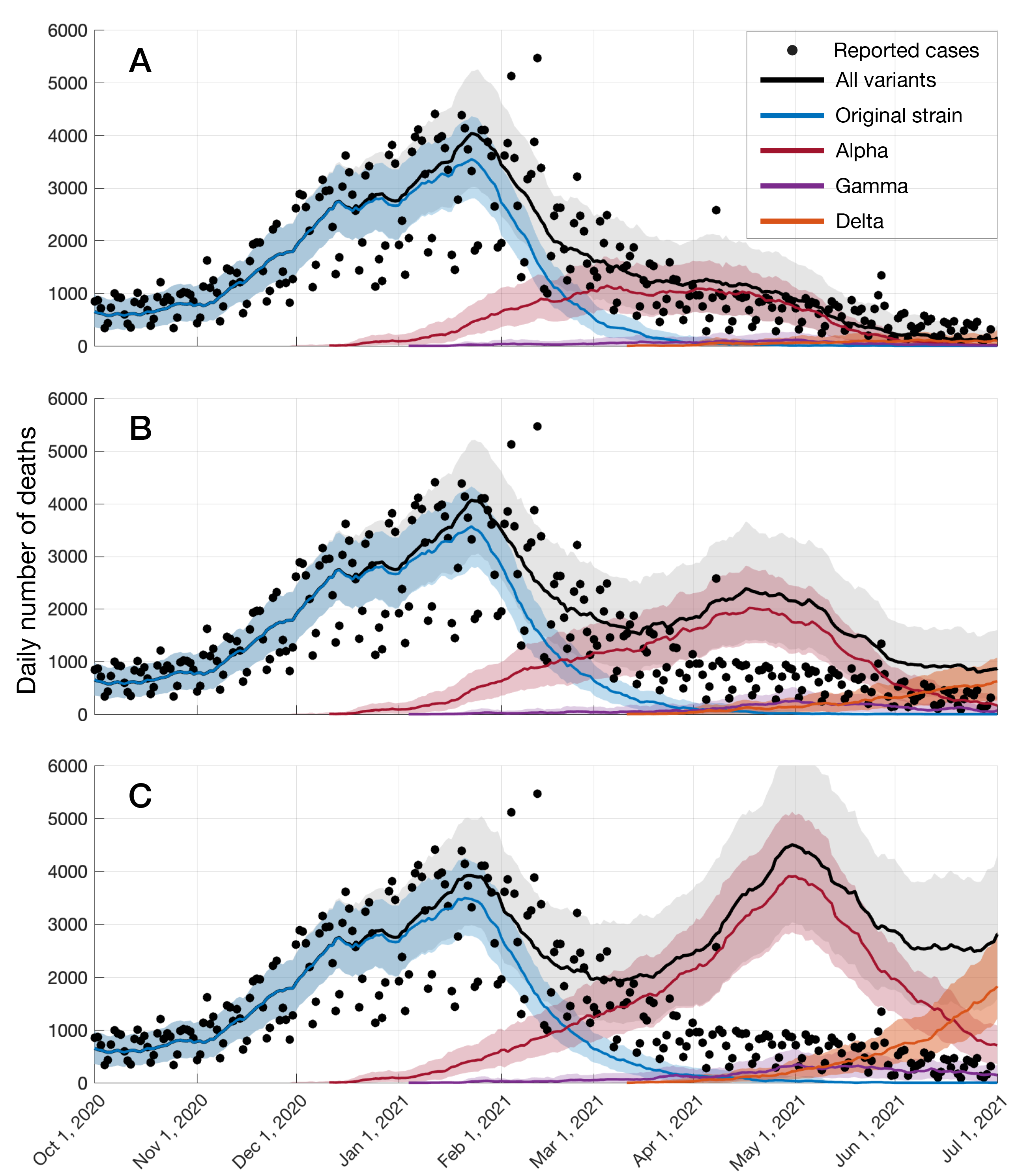
Impact of COVID-19 vaccination progress. Projected number of deaths caused by different variants of SARS-CoV-2 in the US with: (A) vaccination rollout in the US; (B) a temporal vaccination rate reduced to half of the actual pace; and (C) without vaccination. Curves represent overall deaths (black), those attributable to the original variant (blue), Alpha variant (red), Gamma variant (purple), or Delta variant (orange). Solid black dots are reported deaths and shaded regions reflect the range of uncertainty in the projections.

We also found that the swift rollout of the vaccination campaign, which exceeded 3.3 million doses administered per day in April 2021, played a critical role in curtailing COVID-19 spread. Under the second counterfactual scenario (i.e., daily vaccination rates at half the reported pace), we estimated that the US would have endured an additional toll of 120,866 (95% CrI: 93,985 – 146,543) deaths and 457,685 (95% CrI: 351,034 – 570,941) hospitalizations.

The US reported the highest daily COVID-19 cases worldwide for much of 2020 and the beginning of 2021 ^14^. Our results demonstrate the extraordinary impact of swift vaccination for averting cases, hospitalizations and deaths. The speed of vaccination also prevented another potential wave of the outbreak in April that would have otherwise been triggered by the Alpha variant. As new variants – especially the Delta variant – surge among unvaccinated populations this summer, a renewed commitment to vaccine access, particularly for those in historically under-served groups and in counties with low vaccination rates will be crucial to achieving control of the pandemic and preventing avoidable suffering.

## Data Availability

The simulation codes are available at:
https://github.com/thomasvilches/multiple_strains

https://github.com/thomasvilches/multiple_strains

## Acknowledgements

This study was supported by the Commonwealth Fund.

## References

1. ArcGIS Dashboards. Accessed July 2, 2021. https://gisanddata.maps.arcgis.com/apps/dashboards/bda7594740fd40299423467b48e9ecf6

2. The New York Times. See How Vaccinations Are Going in Your County and State. The New York Times. https://www.nytimes.com/interactive/2020/us/covid-19-vaccine-doses.html. Published July 1, 2021. Accessed July 2, 2021.

3. Davies NG, Abbott S, Barnard RC, et al. Estimated transmissibility and impact of SARS-CoV-2 lineage B.1.1.7 in England. Science. 2021;372(6538). doi:10.1126/science.abg3055

4. Challen R, Brooks-Pollock E, Read JM, Dyson L, Tsaneva-Atanasova K, Danon L. Risk of mortality in patients infected with SARS-CoV-2 variant of concern 202012/1: matched cohort study. BMJ. 2021;372:579.

5. Faria NR, Mellan TA, Whittaker C, et al. Genomics and epidemiology of the P.1 SARS-CoV-2 lineage in Manaus, Brazil. Science. 2021;372(6544):815–821.

6. Bolze A, Cirulli ET, Luo S, et al. Rapid displacement of SARS-CoV-2 variant B.1.1.7 by B.1.617.2 and P.1 in the United States. bioRxiv. Published online June 21, 2021. doi:10.1101/2021.06.20.21259195

7. Scientific Advisory Group for Emergencies. SPI-M-O: Consensus statement on COVID-19, 12 May 2021. Published May 14, 2021. Accessed July 6, 2021. https://www.gov.uk/government/publications/spi-m-o-consensus-statement-on-covid-19-12-may-2021

8. Davis C, Logan N, Tyson G, et al. Reduced neutralisation of the Delta (B.1.617.2) SARS-CoV-2 variant of concern following vaccination. doi:10.1101/2021.06.23.21259327

9. Polack FP, Thomas SJ, Kitchin N, et al. Safety and Efficacy of the BNT162b2 mRNA Covid- 19 Vaccine. N Engl J Med. Published online December 10, 2020. doi:10.1056/NEJMoa2034577

10. Baden LR, El Sahly HM, Essink B, et al. Efficacy and Safety of the mRNA-1273 SARS-CoV-2 Vaccine. N Engl J Med. Published online December 30, 2020. doi:10.1056/NEJMoa2035389

11. Sadoff J, Gray G, Vandebosch A, et al. Safety and Efficacy of Single-Dose Ad26.COV2.S Vaccine against Covid-19. N Engl J Med. 2021;384(23):2187–2201.

12. Moghadas SM, Sah P, Vilches TN, Galvani AP. Can the USA return to pre-COVID-19 normal by July 4? Lancet Infect Dis. Published online June 2, 2021. doi:10.1016/S1473-3099(21)00324-8

13. CDC. COVID Data Tracker. Published March 28, 2020. Accessed July 2, 2021. https://covid.cdc.gov/covid-data-tracker/

14. Coronavirus (COVID-19) Cases. Accessed July 2, 2021. https://ourworldindata.org/covid-cases

